# Combined severe-to-profound hearing and vision impairment – experiences of daily life and need of support, an interview study

**DOI:** 10.1101/2023.01.09.23284355

**Authors:** Satu Turunen-Taheri, Annica Hagerman Sirelius, Sten Hellström, Åsa Skjönsberg, Gunnel Backenroth

**Affiliations:** Department of CLINTEC, Division of Ear, Nose and Throat Diseases, Karolinska Institutet, Stockholm, Sweden; Department of Audiology and Neurotology, Karolinska University Hospital, Stockholm, Sweden; Department of CLINTEC, Division of Audiology, Karolinska Institutet, Flemmingsberg, Sweden; Stockholm University, Department of Psychology, Stockholm, Sweden

## Abstract

**Objectives:** The purpose of the present study was to describe experiences of disabilities and factors affecting daily life from the perspective of adult persons with severe-to-profound hearing impairment in combination with severe vision impairment. Furthermore, the study also investigated which kind of support individuals with dual sensory loss received, and their experiences as citizens in the society.

**Methods:** Semi-structured qualitative interviews were performed, analyzed, and categorized using content analysis.

**Results:** Fourteen interviews were performed, with equal number of both sexes. Mean age was 70.1 years (47-81 years). Analysis of the data resulted in 22 categories, six sub-themes and two main themes. Two main themes emerged as *Isolation* and *The Ability to control one’s own daily life*. Surprisingly, most of the participants did not think of their vision and hearing impairment as a combined disability. The interviews showed various kind of strategies to handle daily life. The Deafblind-team unit was reported to offer excellent health care. Companion services for persons with disabilities proved to have become more difficult to get support from and created lack of independence and control over their own lives. However, it was also obvious that the participants felt a positive outlook on life and more solution-oriented in order to adjust their everyday life to their life-situation.

**Conclusions:** The combination of vision and hearing impairment demonstrated isolation, and the respondents in the study have a need of support in everyday lives. At the same time, they struggle to have the ability to control their own lives.

## Introduction

Hearing impairment affects everyday life in communication, and it is associated with health conditions such as social functioning, and mental health [1]. The global burden of hearing loss is a third leading cause of years lived with disability [2]. In Sweden, approximately 18.3%, corresponding to 1.5 million people over 16 years age report hearing loss of varying degrees [3] (estimation: http://www.statistikdatabasen.scb.se/sq/73088). The estimated prevalence of adult patients with severe-to-profound hearing impairment is approximately 0.28%, corresponding to 22.000 of the population in Sweden [4, 5].

The common method to estimate hearing loss is assessing of hearing thresholds with a pure-tone audiometry (PTA) using four frequencies for the pure-tone-average (PTA4: 500, 1000, 2000 and 4000 Hertz) [6]. There are different classifications of hearing impairment and according to the WHO the degree of hearing loss [7] see Figure 1. In 2021, WHO revised the classification and grades of hearing loss, but this study used the previous variant of classification.

**Fig 1.**
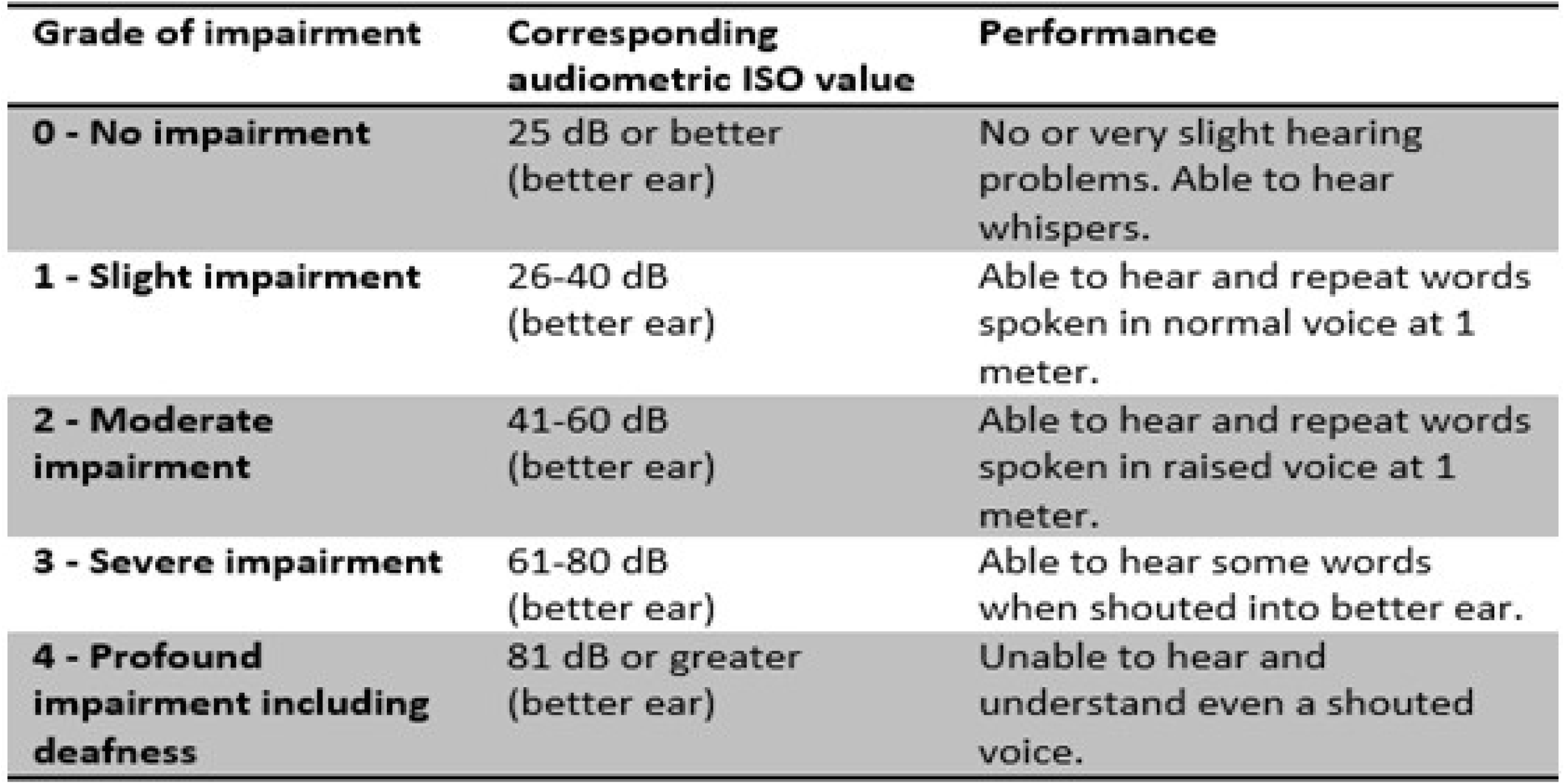
Grades of hearing impairment, World Health Organization (WHO) 1991. Audiometric ISO value = The average of hearing level (HL) at 500, 1000, 2000 and 4000 Hz in the best ear. With permission from WHO to use the table below. Available from: https://apps.who.int/iris/bitstream/handle/10665/58839/WHO_PDH_91.1.pdf?sequence=1&isAllowed=y

Vision impairment has a strong negative impact on daily activities and social participation [8] and increased functional disability [9]. A recent study from Blindness and Vision Impairment Collaborators (GBD) [10] estimates the global prevalence of blindness to 43.3 million people, and moderate to severe vision impairment in 295 million people. Approximately 445.000 people in Sweden has by definition some kind of vision difficulties [3]. However, there is no comprehensive registration of persons with vision impairment in Sweden, but approximately 100.000 people are handled through a Low Vision Clinic, as called ‘Syncentralen’, due to severe vision loss [11].

Vision impairment is defined in four levels: mild, moderate, severe and blindness in the International Classification of Diseases-11 [12] and the classification of vision impairment according to WHO [13] shows in Figure 2.

**Fig 2.**
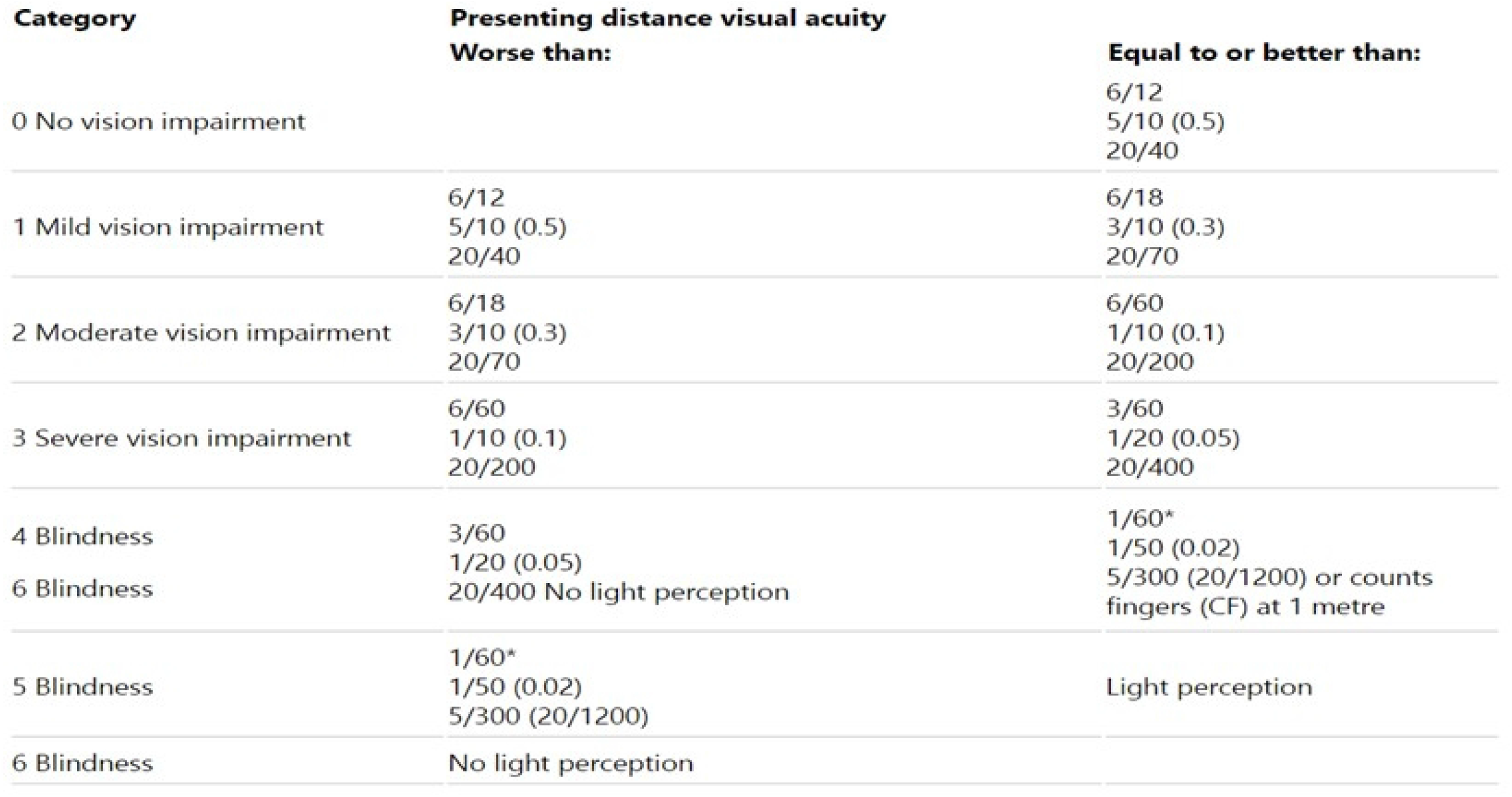
Classification of vision impairment including blindness, World Health Organization (WHO) 2019. With permission from WHO to use the table below. Available from: https://icd.who.int/browse11/l-m/en#/http://id.who.int/icd/entity/1103667651

One disability, such as hearing loss, affects communication and could be associated with reduced quality-of-life (QoL) [14]. A dual sensory loss, i.e., both hearing and vision loss, affects even more everyday functioning and ability to communicate [15]. Globally, estimation of the population of persons living with deafblindness is 0.2-2% [16]. A combined severe-to-profound hearing loss and severe vision loss seems to have more negative impact on patients’ daily life, e.g., anxiety and depression scores are higher compared to severe-to-profound hearing loss alone [17].

The Swedish Association of the Deafblindness [18] states; “Deafblindness is a combined vision and hearing impairment of such severity that it is hard for the impaired senses to compensate for each other. Thus, deafblindness is a distinct disability” [19] (page 1).

In 2006, approximately 1200 people were characterized as deafblind in Sweden [20]. A recently published study [21] shows a prevalence of ∼ 1581 (0.08 %) individuals over 65 years old with a combination of vision and hearing impairment in Sweden. According to another source [22] approximately 2000 people up to 65 years of age are deafblind in Sweden 2018, of which approximately 400 were born deafblind and almost 700 persons are affected by Usher Syndrome. Usher syndrome is the most common cause of deafblindness [23] which is an inherited autosomal recessive disorder with hearing loss at birth and progressive retinitis pigmentosa (RP). The prevalence of Usher has been estimated between 1-4/25.000 people in a worldwide population [24]. In Sweden, the prevalence of Usher for all three clinical types of syndrome is estimated to 3.3/100.000 [25].

Hearing health care organization differs in the arrangement between different regions and county councils in Sweden. Three types of hearing health care practice can be found: traditional “public”, primary authorized “private but publicly funded” and “completely private” [26]. All three types of care exist in Stockholm. Primary authorized clinics take care of patients with mild to moderate hearing loss. Furthermore, the traditional public clinic at Karolinska University Hospital, separately for children and adults, provides care to patients with severe-to-profound hearing loss or combined sensory loss.

Similar to hearing health care, the structure of the vision health care differs between regions and county councils in Sweden. Optician stores can be compared with primary authorized clinics for hearing. The optician refers patient to a doctor if a vision disease is suspected [27], and a doctor can refer patients to a Low Vision Clinic, which offers habilitation and rehabilitation for all age groups of patients with vision loss [11].

In Sweden, specialized care for deafblind persons is offered in the Counselling and Support team for persons with deafblindness, also called “Deafblind-team unit” [28]. They work interdisciplinary and offer a care plan, lectures, group activities, and support in contact with e.g., various authorities, communication training and conversation support, home visit, and training with white cane. The deafblind-team unit also support family members. In Sweden, no referral is required to access the deafblind-team unit, however, it is required that patient has contact with both Low Vision Clinic as well as the unit for extended hearing rehabilitation for adults e.g. at Karolinska University Hospital, and thus fulfils the criteria required for these activities [28].

The Swedish Agency for Participation (MFD) [29] works for people with disabilities to have the right to live an independent life and to be able to participate in society on equal terms as everyone else. Availability is a general principle of the UN Convention on the Rights of Persons with Disabilities [30], both in terms of the physical environment and information and communication. Disability such as deafblindness causes serious consequences that affect the possibility of activity and participation on several levels and when the environment can be adapted to the needs, the possibility of participation increases [31].

Swedish Agency for Accessible Media (MTM) [32], belonging to the administration of the Ministry of Culture, produce and distribute audio books, information, literature and news in easy-to-read Swedish. They make sign language literature accessible and provide support to newspapers transferred to speech.

Patients with hearing loss need an individualized care plan and various types of audiological rehabilitation such as technical rehabilitation and extended audiological rehabilitation. Technical rehabilitation includes hearing aids, cochlear implants, and other technical devices such as connection to smartphones, hearing-loops with telecoil in hearing aid and FM-system. Patients with severe-to-profound hearing loss needs extended audiological rehabilitation with more support with medical, psychological, social and educational care [33]. Backenroth and Ahlner [34] reported on the quality-of-life in a group of persons with moderate to severe hearing loss (n=30; mean age=49.1 years) who had participated in an audiological rehabilitation program at the Department of Audiology, Karolinska Hospital. Participants in the rehabilitation program had learned different coping strategies for their own life situation.

Some of the conclusions of the study were that audiological counselling interventions need to increase the insight in one’s own coping strategies, actively involve family members in the counselling, have more focus on psychosocial working through of emotions and relations, influence the QoL of the hearing-impaired person at the workplace, in short, to have an increased holistic psychosocial perspective to audiological rehabilitation. A recent study by Turunen-Taheri et al [35] showed the importance of extended audiological rehabilitation, group rehabilitation and visits at a hearing rehabilitation educator especially for patients with severe-to-profound hearing impairment.

Similarly, patients with vision impairment need support and efforts with habilitation and rehabilitation that prevents and reduces the difficulties that various forms of vision impairment can bring in daily life. A recent study by Ehn et al [36] shows various life strategies that persons with deafblindness can have by resolving and preventing to handle challenges. Patients need technical devices to manage their everyday lives such as various models of pocket memories, daisy players, magnifying glass, reading TV, reading machine (scans typed text and read it up with a speech synthesis), binoculars, Braille display, visual interpretation, and tactile wristwatch [37].

In Sweden, the Social Services Act (2001:453) [38] ensures that people with disability receive support in chapter 5, in § 7: “The Social Committee shall work to ensure that people who, for physical, psychological or other reasons, encounter significant difficulties in their way of life, have the opportunity to participate in the community of society and to live as others”.

Companion service is for those persons who, because of their disability, needs help to participate in recreational and cultural activities. The purpose is for example to make it possible to meet friends, participate in cultural and community activities and exercise sports activities [39]. In a report from the Swedish National Board of Health and Welfare (Socialstyrelsen) [40] it appears that companion service is an activity that may support people with disabilities to simplify becoming involved in society.

The overall aim of this study was to describe experiences and factors affecting daily life in adult patients with severe-to-profound hearing impairment combined with severe vision impairment. Furthermore, the study also investigates which kind of extended rehabilitation and support patients with dual sensory loss received, and their experiences of being part of the society.

## Materials and methods

The study is designed with a qualitative approach to increase understanding the content [41] by use of semi-structured interviews describing the impact, consequences and experiences of combined severe-to-profound hearing loss and severe vision impairment in daily life, and the support of extended audiological rehabilitation these patients need.

### Method considerations

Qualitative research with interviews aims to describe, explore, and explain the individual participants’ experiences on a deeper level, as for the aims of the present study, experiences and factors affecting daily life in patients with dual sensory loss. To organize and analyse the data collected, the qualitative inductive content analysis were conducted in the study by creating codes, categories and themes [41, 42].

### Selection and inclusion criteria

Recruitment of participants who meet the inclusion criteria: severe-to-profound hearing loss at 61 dB HL or more in the best ear [7] in combination with severe vision impairment, were performed in 2018-19 in consecutive patients with self-selection. Patients were seeking care at the County Council in Stockholm at the Department of Audiology and Neurotology, Extended Hearing Rehabilitation, Rosenlund, Karolinska University Hospital, Stockholm and at the Counselling and Support team for persons with deafblindness so called the Deafblind-team unit, at Sabbatsbergs Hospital, Stockholm, Sweden.

To be included in the present study, patients had severe vision impairment to blindness, and severe-to-profound hearing impairment according to WHO previous grades of hearing impairment (Figure 1). Even patients depending on a sign language or tactile sign language interpreter were included.

The researchers conducting the interviews informed their colleagues at the units about the study and were asked to recruit patients to participate in the study. The authors did not contact the patients themselves in order to not influence their decisions to participate in the study.

The study included 14 patients from Stockholm County council. Seven males and seven females participated in the interviews. The age ranged between 48 to 81 (mean 70.1) years (Table 1). Seven of the participants were recruited through Karolinska University Hospital and seven through Sabbatsbergs Hospital. All participants had participated in audiological rehabilitation at the Department of Audiology and Neurotology, Extended Hearing Rehabilitation Rosenlund, Karolinska University Hospital.

**Table 1.** Demographic data. Gender, age, hearing and vision loss distribution of the interview patients

| Characteristics |  |  |  |
| --- | --- | --- | --- |
| Gender | Women | Men | Total |
| Age |  |  |  |
| 41-50 | 1 | 1 | 2 |
| 51-60 | 1 |  | 1 |
| 61-70 |  | 3 | 3 |
| 71-80 | 4 | 1 | 5 |
| 81-90 | 1 | 2 | 3 |
| Total | 7 | 7 | 14 |
| Mean age (year) | 70.8 | 69.4 | 70.1 |
| Hearing loss PTA4<sup>1</sup><br>best ear, mean (range) | 69.4 (60-95) | 75.1 (68-91) | 72.2 (60-95) |
| Vision loss, severe<sup>2</sup> | 3 | 3 | 6 |
| Blindness, total | 4<sup>3</sup> | 4<sup>4</sup> | 8 |

<sup>1</sup> Hearing loss PTA4 means the average of hearing level (HL) at 500, 1000, 2000 and 4000 Hz in the best ear.
<sup>2</sup> Severe vision loss as, for example, caused by Usher II, retinitis pigmentosa, glaucoma, keratoconus, macular degeneration.
<sup>3</sup> One participant is blind at one eye and severe vision loss in the other eye. One participant can only see dark and light.
<sup>4</sup> One participant is blind at one eye and severe vision loss in the other eye.

### Data collection

Face-to-face interviews were conducted in Swedish by two of the authors (STT and AHS) at the unit of Extended Hearing Rehabilitation, unless the patient wanted to meet in another place, such as the home environment or their workplace. Thirteen interviews were performed at the hospital and the remaining one at the patient’s workplace. A semi-structured interview guide (Supplementary material) was used but participants were also given an opportunity for deeper discussions and encouraged to open dialogue. Prior to the investigation, the interview guide was pre-tested on a test interview with one patient having moderate to severe hearing loss and severe vision loss. The pre-interview led to that some added questions. Both interviewers (STT and AHS) participated in each interview, one as the active conversation leader and the other observed the interview and wrote field notes. The interviewer and the observer switched the roles every second interview section. Interviews were audio recorded with a digital Dictaphone.

The interviews were performed in a quiet room with the possibility for the participants to listen by a hearing-loop while the interviewer talked in a microphone. One of the interview sessions did not have access to t-loop due to technical problems. The interview at the participant’s workplace was performed in a coffee room. In three of the interview sessions, a companion service person participated in the interview session, and one of those was a relative. No interpreter was present in any of the interviews. The authors were allowed to record the interviews on audio recorders.

All interviews were transcribed verbatim in Swedish, which means that all verbal and nonverbal [e.g., coughs] utterances and pauses were transcribed. The interview times lasted between 41-120 minutes, totally 995 minutes for all 14 interviews. Transcription was done by a person outside the study. Both authors STT and AHS listened through the recordings afterwards. The transcribed text was read through several times by two authors (STT and AHS). For quality control purpose, one of the authors (ÅS) listened to all interviews and took ongoing notes of the interviews. The authors (GB and SH) controlled all transcribed texts. Each participant received their own transcribed text by mail or email.

### Data analysis

Interviews were analysed and categorized using content analysis. Content analysis is defined as “a research technique for making replicable and valid inferences from texts (or other meaningful matter) to the contexts of their use” [42] (page 24). The present study used inductive content analysis describing interview areas with open coding [43], formulating codes with condensed meaning units [44], analysing texts into categories [42], sub-categories, codes and themes [44, 45].

The process was performed in phases. The written data and audio material was listened to and read through several times. The content analysis was performed at first by two of the authors (STT and AHS) together, and thereafter with four of the authors (STT, AHS, ÅS and GB) together. The fifth author (SH) has read the transcribed texts. The analysis was discussed among the researchers. The meaning units were identified to codes and categories. Coding was regularly reviewed by the research team (STT, AHS, ÅS and GB) to identify and incorporate emergent themes. The data were then examined by author STT and author AHS to identify typical quotes and common reasoning.

The selected quotes texts for manuscript were translated from Swedish to English. For the reader’s sake, the translated texts have been grammatically written in English and we used “text […] text” if any sentence were uncompleted.

To consider transferability, trustworthiness, credibility, reflexivity, and confirmability, four authors (STT, AHS, ÅS and GB) checked the elaborated findings for relevance while carefully considering the meaning units and clustering before finalising the categorisation and themes [46-48].

The authors followed the phases of Braun and Clarke [49], and also creating categories as many researches describe for inductive content analysis [43, 45] see revised model Figure 3. The authors have followed gold standards [50] and the consolidated criteria for reporting qualitative research (COREQ) [51]. The text was coded and categorized to find meaningful structures. Participants were named as P1-P14. Every code were labelled with the individual marking by P1:1:1 or P1:1:a:1 which signifies an own identity for each quote. First digit is participant’s number, second digit describes category, third digit is for the quotation’s serial number, and an eventual small letter corresponds to a sub-category. All authors evaluated codes, categories and sub-themes and theme to obtain the consensus.

**Fig 3.**
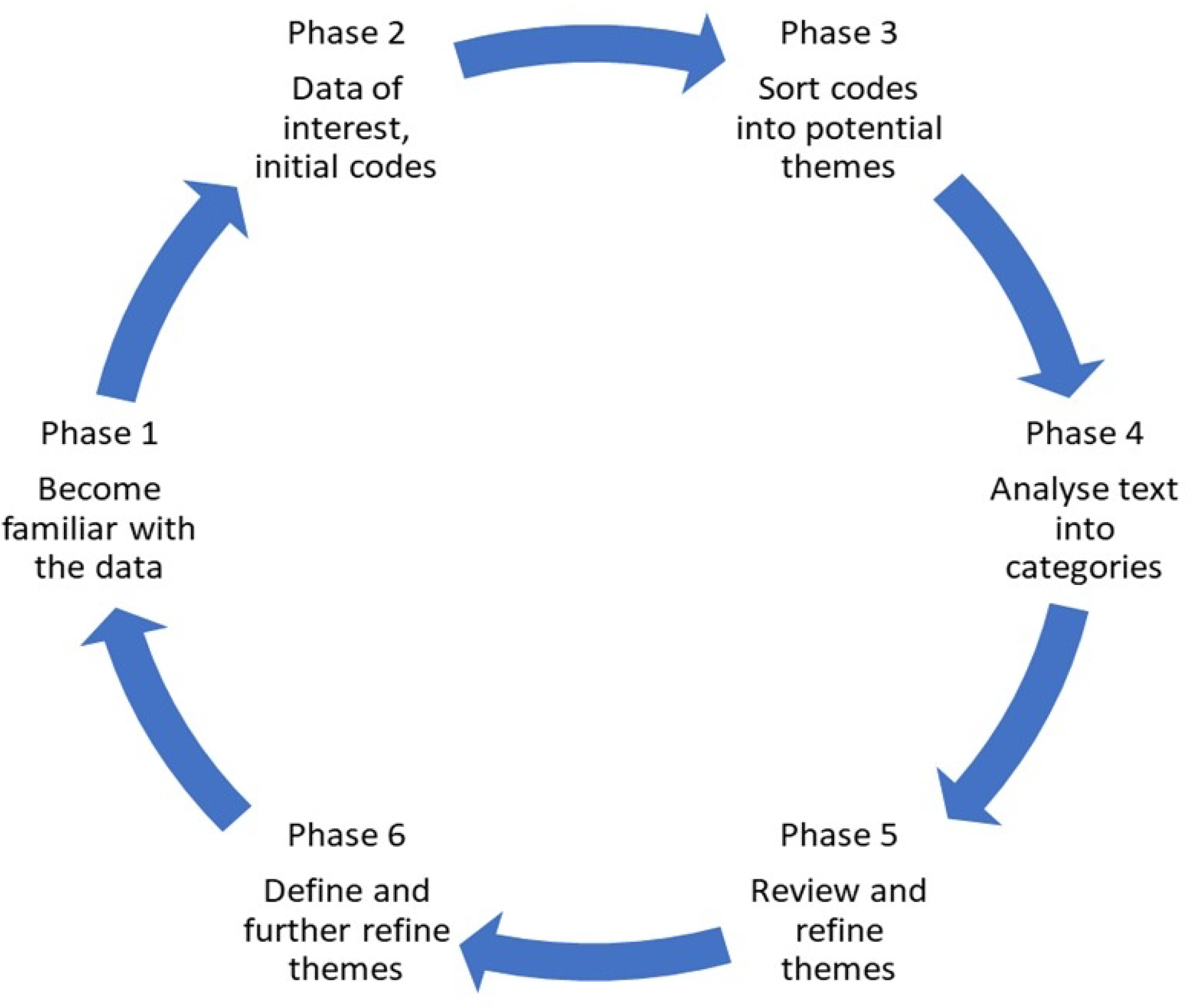
Phases of data analysis in the study. Revised model in the present study based on Braun and Clarke analysis (2006).

### Ethical considerations

This study was approved by the Regional Ethical Review Board, Stockholm (2014/2101-31). All patients were provided with both written and verbal information before they were included in the study.

According to the ethical guidelines, the original ethical consent did not include the question of citation use, why participants afterwards were asked for the consent that applies to the use of quotes from the interview. The quotes were anonymous so they cannot be coupled to a specific individual. Three participants refrained from accepting the consent for the use of their quotes, hence their responses have been deleted from the quotes.

## Results

### Demographic data

In total, 14 interviews were performed, with equal numbers from both sexes. The data analysis resulted in 20 categories, six (6) sub-themes and two (2) main themes that are presented below. The abstraction of categories, sub-themes, and the main themes are shown in Table 2 and Figure 4.

**Table 2.** Themes, sub-themes, and categories based on conversations areas.

| **Themes** | **Sub-themes** | **Categories** |
| --- | --- | --- |
| Isolation<br>The ability to control one's own daily life | Contact arenas | ○ Professional network (Hearing Health Care, Low Vision Clinic, Deafblind-team unit)<br>○ Family network<br>○ Daily social network |
|  | Time perspective | ○ Life history<br>○ Current situation<br>○ Future |
|  | Disability | ○ The Combinations's effects (hearing and vision loss)<br>○ Possibilities<br>○ Limitations |
|  | Challenges | ○ Communication barriers<br>○ Information to the environment<br>○ Demands |
|  |  | ○ Emotions<br>○ Self-image<br>○ Social competence |
|  | Needs | ○ Continuity in professional network<br>○ Availability<br>○ Technical tools (including White cane)<br>○ Companion service |
|  | Coping strategies | ○ Cognitive<br>○ Emotional<br>○ Problem focused |

**Fig 4.**
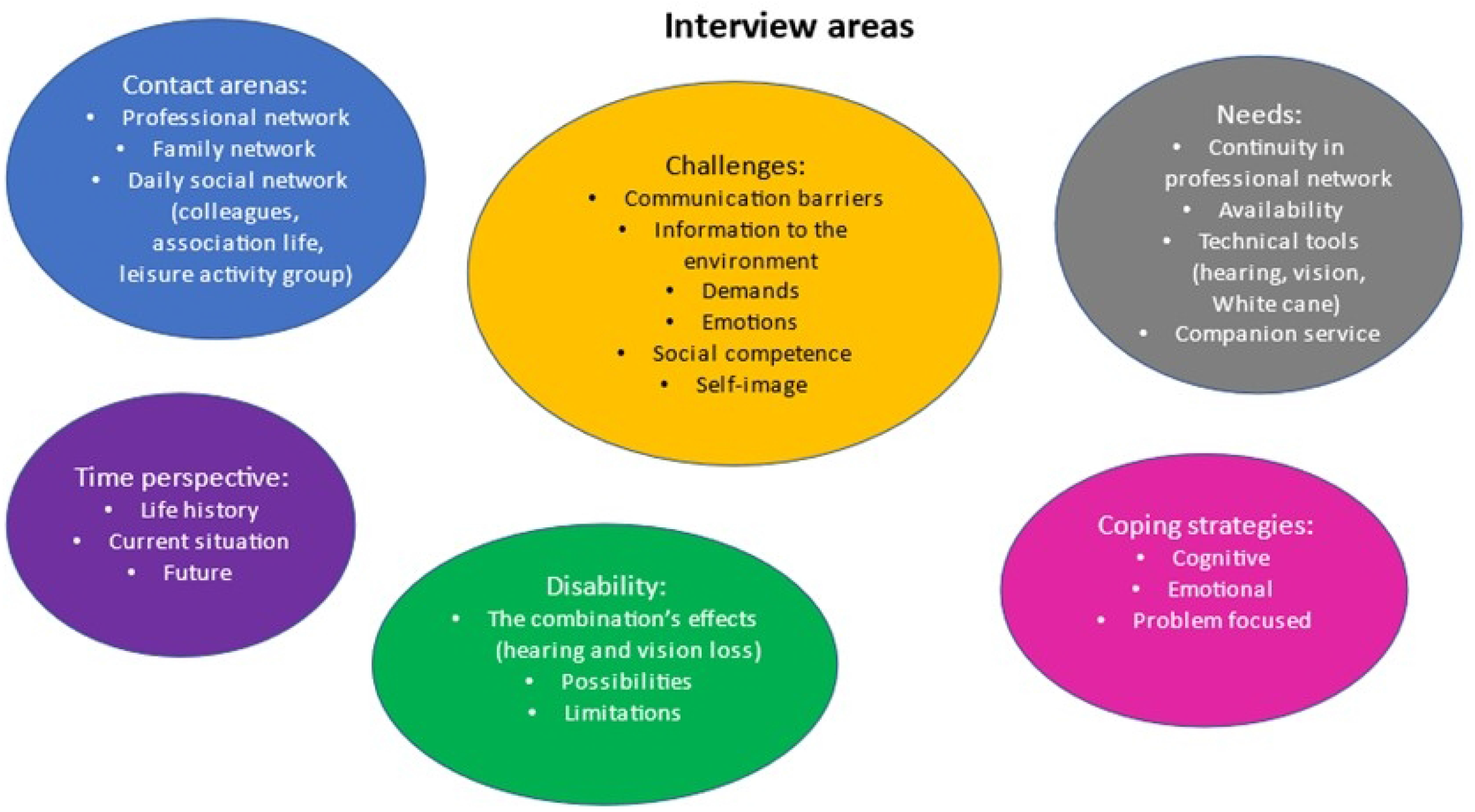
Interview areas in the study resulted in six sub-themes and 20 categories. The abstraction of sub-themes and categories.

Causes of hearing loss for this study population were e.g., noise trauma, hearing loss since birth and/or childhood, Meniere’s disease, Usher Syndrome. The average of hearing thresholds was PTA4 for right ear 75.21 dB HL and for left ear 76.00 dB HL. PTA4, *best* ear, for females were 69.42 dB HL and for males 75.14 dB HL (Table 1).

Causes of vision impairment for the study population varied and were Usher Syndrome, keratoconus, retinitis pigmentosa, external damage to the eyes in childhood, congenital blindness, glaucoma, rheumatism in childhood, incubator treatment at birth, and macular degeneration. Six participants had severe vision loss and eight was totally blind (two of them in one eye and with severe vision loss in the other eye) (Table 1).

In three cases, the hearing impairment was the first impairment from birth and in three cases the vision impairment existed from birth, and the remaining eight participants acquired disabilities in childhood or later in life.

### The main themes

The qualitative content analysis of the categories and sub-themes formed in two main themes “*Isolation*” and “*The ability to control one’s own daily life*”. These main themes emerged from the sex sub-themes presented below (Table 2).

### Sub-themes

Six sub-themes: *Contact arenas, Time perspective, Disability, Challenges, Needs, Coping strategies*, emerged from 22 categories.

### Categories

The following 22 categories were identified; Professional network (such as Hearing Health Care, Low Vision Centre, Deafblind-team unit), Family network, Daily social network (such as colleagues, association life, leisure activity groups), Life history, Current situation, Future, the Combination’s effects (hearing and vision loss), Possibilities, Limitations, Communication barriers (such as Others (mis)understanding/attitudes, Own (mis)understanding/attitudes), Information to the environment, Demands, Emotions, Self-image, Social competence, Continuity in professional network, Availability, Technical tools (such as for hearing and vision impairment, and separately White cane), Companion service, and Cognitive, Emotional and Problem focused strategies (Figure 4, Table 2).

Here follows a summary of the sub-themes that emerged to the two main themes.

### Sub-theme: Contact arenas

The categories *Professional network (including Hearing Health Care, Low Vision Clinic and Deafblind-team unit), Related persons and Colleagues* represent different **Contact arenas** the participants have.

In general, the participants were satisfied with the care they have received and the support that the medical team provided. They were properly taken care of.

However, participants could comment on *the Hearing Health Care* that:

> “… *but it feels somehow that, uh, one is using county council’s funds, money and resources…”* (P1, male)
>
> *“Yes I know Hearing health care exist and it has been great and so with the Low Vision Clinic as well. A great support and help in my experience”* (P12, female)
>
> *“Eh, yes there has been more changes in staff lately. Well, then one was unsure and wondered which audiologist one will meet… I think it has been working well and fells under control and I have felt safe”* (P14, male)

There are some re-occurring issues in need for improvement. Some based on very long waiting time to get an appointment and some logistical issues. There was also a request for more time needed to explain the use of all technical aids. Most participants commented on the long waiting time before receiving care at *the Low Vision Clinic* at which there is also a need of more employees.

> “*Yes, it is rather positive. Eh I think well, it is like the same there, they are hard to get appointment at * laugh *, it takes time. Very long time”* (P1, male)
>
> *“It’s a little tricky to reach there, or up to that floor. There are many levels* … *so it is good most of the times, but it takes an incredibly long time between appointments”* (P2, female)
>
> *“*… *lack of personnel. That leads to very long waits* … *Yes, it is more staff. That’s what they need. That is the most important thing”* (P6, male)
>
> *“Visiting hours is too short. They give people technical aids, now I don’t talk about myself, and people go home with them in boxes and haven’t a clue how to use them”* (P10, female)

All participants only had positive statements to say about *the Deafblind-team unit*, at which patients felt involved in their care, staff owned high-level of competence and a special understanding of the patient and their needs. Participants also felt they were well informed and involved in their own care which is very important to create trust. They felt they were met with care and always understanding by all staff members:

> *“And eh the Deafblind-team unit serves as my second home* … *I got a new life after I got in touch with them”* (P4, female)
>
> *“From the deafblind-team unit the effort is excellent”* (P5, female)
>
> *“Yes they have the knowledge* … *they have understanding* … *One gets a lot of empathy”* (P14, male)

The importance of *Family network* is shown in several quotes. It is emerged that sometimes participants had to remind relatives that they hear and/or see poorly, and in some cases, the closest ones could be the ones who showed the least understanding of all.

> *“… for me it is well arranged because I have a husband who helps a lot at home such as vision interpretation and all that with document and finances and, uh, and that I do ask about food recipes and so on”* (P2, female)
>
> *“Then I need to burden my friends, but it works, it works to a certain limit”* (P6, male)
>
> *“*… *the world is starting to fall apart. Mhm. So I don’t have the network I want”* (P10, female)

The category *Daily social network* such as *colleagues* shows that workplace adaptations are required for a person with vision and hearing impairment. This can apply, for example, marks where stairs are located, and contrasts with colour to make it easier to see differences of levels. It is important that colleagues in the workplace address themselves using their name when they talk to someone with a severe vision impairment in order to rule out the insecurity that can occur when you don’t know who you are talking to.

> “*New environments are really hard for me, eh, but then I have my work colleagues, they help me all the time* … *Or when I come into the copy machine room, there is also a person saying ‘hello P5, this is M…’’* (P5, female)

### Sub-theme: Time perspective

The categories about *Life history, Current situation, and Future* formed sub-theme **Time perspective**.

Many participants perceived *Future* as more threatening rather than safe. The future appears unhopeful, depressing, isolating, with anxiety. Some participants wish for progress in ophthalmic surgery. It could for example be manifested by being afraid of getting a much-worsened hearing or vision impairment.

> *“Yes, is there any future for me in my situation?* … *It feels hopeless”* (P4, female)
>
> *“No, but it is, progress is also being made when it comes to eye surgery. It is my hope, as I said, perhaps I can get a little better vision in the future”* (P6, male)
>
> *“I have accepted that I suffer from this… But for me, I have the premise that I’m grateful every morning I wake up and I’m grateful that I still have some vision left”* (P9, male)
>
> *“Yes, I think so because it is downright depressing. Accordingly, it is isolating”* (P10, female)
>
> *“Yes, of course one hopes that one will get the vision back, but it is clear that it will not happen”* (P13, male)
>
> *“Yes, I struggle with a lot of anxieties and worries *pause* I have not much positive thoughts…one rather take one day at the time and seize the day”* (P14, male)

### Sub-theme: Disability

*The Combination’s effects* by a simultaneous hearing and vision loss, *Possibilities*, and *Limitations* formed sub-theme **Disability**. Most participants had not realized that they have a combination of disabilities, more likely they thought of two separate disabilities independent of each other. Some participants with congenital blindness, emphasized the great importance of hearing that remained. Some participants, on the contrary, had the experience that it was the vision loss that was the cause of the isolation. One participant mentioned that not until he/she no longer could read from other people’s lips or see the body language, he/she discovered that he/she suffers from a combination of disabilities. The *combination’s effects* of disabilities limit and isolates the independence, for example, to go out without a companion:

> *“No, but that is that I have not thought about it as much as a combination so that sometimes it is difficult from the vision point of view, and sometimes it is difficult from a hearing point of view and eh I have not thought of it clearly that it is just the combination that makes it extra difficult* … *then because I have not thought so much about it that I both see and hear poorly and I have seen more that sometimes there are vision problems and in other situations hearing problems”* (P1, male)
>
> *“For the vision impairment, I had learned how to deal with… then this second thing came and made trouble. Because if you are ‘only’ vision impaired then you are using the hearing to locate yourself* … *Yes, it is not a fun combination!”* (P4, female)

Many participants mentioned difficulties such as ability to orient oneself and move in various environments.

> *“It isolates me. In particular, if I saw a little earlier, I could at least go out on myself. Because, then hearing was not an obstacle. It was not. But with the combination then, I am isolated in that way. I depend on people constantly to be able to come outdoors.”* (P10, female)
>
> *“Yes. It is not easy to adapt, I think. That, it has decreased gradually, you may say with the hearing as well…. it is the loss of hearing that has the greatest impact, leading so that I need more help really”* (P12, female)
>
> *“Yes, unfortunately I have become more isolated because of the vision impairment now during the last 15 years. And it has also made me more frustrated and irritated. For example, you stumble upon things on… and you need more support… but even there when you have difficulties, you can no longer use lip reading. It makes me feel the need to have a good hearing aid. But again, this means that you become more isolated, you are better off and safer at your own home”* (P14, male)

Some participants may see their vision and hearing impairment as an asset as, for example, becoming more detailed in what they are doing or that another sense can be strengthened as a resource in *Possibilities*:

> “*I guess I never…* .*thought of that oh, now I can not do this because I both see and hear badly. Of course, there are things I can not do because of this combination of disabilities. But eh, it’s hard to figure out what it would be *laughs*”* (P1, male)
>
> *“But I’m thinking of ‘Yes, this is as usual. This is what I’ll do’ and then I stand there and ‘“Oh, I do not hear this, I do not see this, but I will participate”*. (P4, female)
>
> “*I am very careful and because one has such a limited field of vision it leads to that I do not miss anything usually *laugh*. I am very neatly, and it takes some time but I am very careful and systematic…”* (P5, female)
>
> “*Because when we meet through various associations. Then I meet a lot of people but then I have an interpreter. Whom I book. This person is then my eyes and ears*” (P14, male)

The participants reported a lot about *Limitations* in their lives. There were difficulties in being able to move and orientate oneself, being in danger in traffic, not being able to shop, not being able to read, not being able to drive a car, being dependent on other people, and not have the freedom to do what you want. Some have been able to take the municipal subway or bus in the past, but don’t dare to do so anymore. Public transport has many physical and social barriers to overcome such as platform gaps, inadequate travel information and noise. Here are some of many limitations that are addressed:

> *“Other changes concern of course the daily newspaper reading eh is not at all in the same way, it is a big difference, even though my wife then reads for me, it becomes limited due to what she thinks is interesting then* …*”* (P1, male)
>
> *“And it’s really very important to keep active, uh, because you can’t move as much as others when you don’t see* … *And of course, it’s difficult to communicate and hear them at the same time. Uh, so you feel very, very lonely in your surroundings* … *you are so dependent to have a person on your side, one’s side, in many situations* …*”* (P2, female)
>
> *“The combination only causes problems”* (P4, female)
>
> *“How does it affect my everyday life? Continuously”* (P5, female)
>
> *“This being able to read, it’s tricky, I can’t do it. I don’t even see what the names of the magazine are* … *Oh, so we run through the week’s mail and such. And bills and things like that”* (P6, male)

Many raised the need for companion service and commented upon the limitations and restrictions of this service being reduced in time. Some participants mentioned the lack of feeling of freedom to be able to do things they wanted without the help of others. Most participants address the lack of being able to go and buy their food all by themselves.

> *“*… *But then it was good to have companion service too* … *But the limitations are notable of course, yes. That you do not have that independence to do things spontaneously”* (P9, male)
>
> *“There are limitations when you can’t go out on your own. But, I need help, that is probably the biggest problem* … *What is important to me, is actually to have companion service. And I get that. However, I need more companionship hours”* (P12, female)
>
> *“… When I go shopping myself. Because it is a gorgeous feeling, a freedom that, if you are thinking like a toddler, ‘I can’. And for me it is important to try out, when I come to a grocery store and I go there and pick for myself…”* (P14, male)

### Sub-theme: Challenges

Sub-theme **Challenges** emerged from six (6) categories:

Category *Communication barriers* demonstrated the importance of introducing oneself every time encountering a person who has severe vision impairment or is blind to avoid misunderstanding.

> *“So it’s always, uh, I’d like to get an introduction every time I meet a person*.
>
> *It’s so sad that you never know whom you meet unless the person says, ‘Now comes Karin’ or ‘Now comes that one and the other*.*’”* (P2, female)
>
> *“… Then we met a couple who stopped by and cheered and sounded so familiar, but I hadn’t, I couldn’t figure out whom it could be. And eh, I had then introduced myself well, then I asked my husband ‘But with whom did we talk to?’ Then it turned out that it was a couple that we had known for 40 years*.*”* (P4, female)
>
> *“*… *when I don’t hear what they say. Then they get tired of talking to me. Then they turn to the companion service person. Or whoever is with me. So the situation can be so crazy and I feel so offended”* (P10, female)

There is a lack of understanding in communication with other persons, or neighbours leading to others or own (mis)understanding/attitudes:

> *“Oh my husband sat opposite me and I wanted to talk towards him so I used sign language, and then the lady sitting next to me says ‘I don’t like you using the sign language because I get the feeling that you’re talking about me’“* (P4, female)
>
> *“Yeah, I felt like… I have always been told that the hearing impaired has a suspicious mind, when not hearing what others are talking about then you think they are talking about you. But I have never thought that even others can be suspicious”* (P4, female)

Category *Information to the environment* illustrates a relatively new way of informing that a person suffers from combined vision and hearing impairment – the use of a white cane with red markings.

> *“*… *if I do not say hello to you, it is not because I am impolite, it is because I do not see you, so come up to me and say hello or pat on me. So, you can say hello”* (P5, female)

These disabilities cause fatigue and takes energy in many ways forming a sub-theme *Demands*:

> *“Well, they don’t understand… that e, they say you are after all as usual. But you have no idea how much it costs to try to be as usual. It takes almost all energy* … *So, I am always on tenterhooks. Oh, that also takes a lot of energy*.*”* (P4, female)
>
> *“…I had not energy enough to read a fairy tale to my children* … *I had constant pain throughout my whole body. I had a headache, I had pain in my eyes but in some way, it was the daily grid, this was ongoing, ongoing, how can I express myself* … *That one was in pain all the time”* (P5, female)

Perceived sadness and sorrows could appear when participants cannot ski any longer or drive a car, which was discovered also in the category *Emotions*. Many participants had fears with a concern of decline in hearing or vision, and some mentioned to be scared near children not to hurt:

> *So yes, regarding children of course. I never dare to go near to children because I am so scared to step on them, hm, yes* … *So, I am also very scared of snow, uh, nowadays, although I used to like snow, skiing, and stuff like that before. Now I don’t like it at all”* (P2, female)
>
> *“Aa, but I hope that it will not worsen in any case with the vision so what I have left, will remain”* (P13, male)
>
> *“So that, we really miss this very much that maybe not*… *keeping the car longer, when we had, we travelled a lot and looked around everywhere and I really do miss that*.*”* (P13, male)

In category *Social competence* were social skills described as very important to be able to move on in life and to get it in the best way is by spending time with other people.

Category *Self-image* shows that many struggles enormously, and they do not give up easily but want to prove that they can still manage their everyday life, even in a different way. One participant mentioned that he/she does not give up but wants to show that everything is possible for his/her own survival and to be a role model for others. Another participant wants to fend for himself as much as possible:

> *“*… *I think most people do so. But I want to try to do as much as I can myself. It’s a bit stubborn there too but…*laugh*”* (P9, male)

### Sub-theme: Needs

Four categories appeared to sub-theme **Needs** which reflects different basic needs in daily life. Category *Continuity in professional network* showed the desire to be called when it is time to change hearing aids, just as the deafblind team invited their patients every year:

> *“*… *for the Deafblind-team unit are somewhat a unique in that manner that they every year are calling you for an appointment to discuss the rehabilitation plan”* (P9, male)

Information regarding society issues takes place to a large extent today through TV, radio, internet, social media, mobile phones. This demands good vision and hearing. In the category *Availability* appeared many difficulties related to lack of the possibility to participate through media. This may lead to isolation if one does not get accessibility in society. Radio is perceived by many as a very good source of information.

> *“Much is made for normal sighted people and normal hearing people”* (P2, female)
>
> *“A lot is obtained by computer”* (P4, female)
>
> *“… However, I know that with social medias it has been an incredible upswing. I get a lot more information today than I got a few years ago* … *Eh, but, in particular social media, make me keep up with today’s news and global news in a different way than I did before”* (P5, female)
>
> *“The radio is probably presenting the best global news media channel … as you get information about the whole world”* (P6, male)
>
> *“It is then through text TV. And eh internet. Eh, I have trouble to get information through the mailbox* … *this with paper, it is, of course, and it is, you feel you hate to bother with paper…to connect with surrounding world…Yes then you really get isolated”* (P14, male)

Participants have received various kinds of *Technical tools* for both vision and hearing impairment in their rehabilitation. Some quotes show about these aids:

> “…*speech syntheses can only be, they cannot be perfect but you have to interpret slightly what they say* …*”* (P1, male)
>
> *“…Yes, about walking around with a cane, it is also a circumstance that you have learned how to get along with cane”* (P2, female)
>
> *“And also, this with wind noise and, then you get very tired and irritated”* (P14, male)

In the present study, the *White cane* seems to have a special important place but also showed the vulnerability with the cane. Many participants describe that it took time before they became “friends” with their cane and that it required cane training for the technique. Some were ashamed in the beginning, someone was afraid to announce that you can rob her/him, but most of them mentioned the white cane as their most important aid.

> *“But it was a big step from getting the cane to using it. It took me many years*. *… Oh, we – me and my child - meet an elderly lady who stopped right in front of us, just stood staring. Oh, then my child says ‘What does that aunt look at Mommy?’ ‘She’s probably looking at my cane’ I said. “Then you have to choose, either me or the cane,” the kid said. What do you choose? So, then it was* … *Then I stopped using the cane. Because he/she was probably ashamed, or thought it was embarrassing… Now the cane is my most loyal companion”* (P4, female)
>
> *“I mainly use this white cane in unknown surroundings or is walking in unknown surroundings, in the dark”* (P8, female)
>
> *“Eh, a little sporadic at first, but now in these last years I always take my cane with me when I am leaving home. I think it’s very good* … *No, you should never underestimate that white cane. It has a great significance*.*”* (P9, male)
>
> *“And then now if I go outside and wait if I don’t have the rollator* … *then I don’t fold my white cane at all to signal that here I stand and don’t see. And that is because I do not want people to feel that I am an easy catch*.*”* (P10, female)

The *Companion service* represents the important need for support for persons with dual sensory loss. This service proves to be a very important part of vision and hearing disabled lives. Eight of the 14 patients had support by the companion service in varying degrees. Two had companion service one (1) hour a week (4 hours per month), one had five hours per month, one had 10 hours per month, two had 20 hours per month, one had 30 hours per month, and one had 40 hours per month. Six of the participants had no companion service at all at the time of the interviews, of whom at least four had closely related persons as support. Many participants mention a problem with the fact that lately the companion service hours have decreased compared to previously.

> *“Uh, then I can add that now, I have companion service, 20 hours per month. They however reduced the hours, so I have been mentally cracked for a year…*.*That’s it, and I’m not afraid to be addicted, I’m used to it. But there are limits to how much help I can get”* (P10, female)
>
> *“It is the companion service that is important in fact, and these 20 hours are not always enough”* (P12, female)
>
> “*The companion service is 5 hours a week…*.*I try to take it coherent so we have time together do something substantial. Companion service adds more quality if you can get more coherent time together”* (P14, male)

### Sub-theme: Coping strategies

The last categories; *Cognitive, Emotional and Problem focused strategies* emerged to sub-theme **Coping strategies**

Participants in the study have various *Strategies* showing resourcefulness and conformability. Several participants expressed that hearing and vision disabilities resulted in various strategies. The strategi may be to strive for to try to live as normal as possible by using problem focused coping strategies. One participant shared that he/she constantly saw opportunities and adapted to his/her problems. Some participants used their creativity and ingenuity in outdoors and sometimes a lot of planning was required the day before to be able to move to new places.

> *“Yes, no, so I am not very shy, but when I think I need to solve a problem and there are some people nearby who seem to be able to help me, then I ask* … *I find that one get very good help many times of fellow human beings”* (P1, male)
>
> *“*… *a very good way to get through a forest is with support of a rope, a short, uh, yes, one (1) meter of rope, which is fixed to the companion’s backpack in front of you, for example and then I have one end, and then we go with walking-sticks. Then I have the rope in one hand, uh, and make sure the rope is stretched, because as soon as it goes down, I stop, because otherwise you will collide “* (P2, female)
>
> *“I don’t run to the bus, I walk, and if I miss it, I take the next one, I don’t want to stumble in my life. In contrast, I take it easy and safe*.*”* (P5, female)
>
> *“Well then I have to be there a long time in advance to be able to find that address I am looking for. And sometimes, if it is close then I go there the day before to see how it looks like”* (P8, female)
>
> *“Of course, I try to go shopping in the mornings when there are limited people out there* … *I only go to the gym during daytime, it would never occur to me to go there on weekends or evenings of course”* (P9, male)
>
> *“*… *then I usually ask if it is okey that I go along with a person”* (P14, male)

## Discussion

The present study is performed with a qualitative methodology with aspects of daily life and need of support from the perspective of participants with dual sensory loss which is not common in this kind of research. As reported in Backenroth & Ahlner [34] studies with a qualitative approach in audiological research has first been reported in the late 1990’s.

The aim of the present study was to describe experiences of disabilities and factors affecting daily life with dual sensory loss. The content analysis resulted in 22 categories and six sub-themes. During processing the categories, two main themes emerged as *Isolation* and *Ability to control one’s own daily life*.

Many participants experienced the lack of control and independence over their own lives. Isolation appeared in various categories.

Interestingly, the participants, during these interviews, realised that they suffered a combination of sensory loss and not just two separate disabilities.

### Result discussion

Participants in the study had suffered from their disabilities for various lengths of time, some were born with vision or hearing impairment, and some acquired later in life.

The present study had an even number of participants in terms of gender, but a wide range of participants’ demographics, such as types of hearing loss and visual impairment, and causes of the combination. Previous national studies in this area have often examined patients with combined hearing and vision disabilities in patients with solely Usher syndrome [25, 52, 53]. In the present study only four (4) out of 14 suffered from Usher syndrome.

In sub-theme *Contact arenas*, the Deafblind-team unit demonstrates to deliver an excellent and successful care. The Deafblind-team unit handles with many dimensions, not only with the individual but with the whole family. They work individually, in groups and in teams, interprofessional with hearing and vision educators, counsellors, psychologists and consulted doctors. They have continuity when they call patients, and all patients have two contact persons at the Deafblind-team unit. They offer e.g. conversation groups with older couples, sign language, stress management, fitness group, white cane training, and cooking group.

The present study emphasizes the importance of having related persons and friends since you are dependent on another person to read mail and newspaper, manage your finances, visit a doctor, go for a walk, skiing, swimming, or shopping, take a bus and subway, or drive a car as some of them could do before. Many mentioned mail readings as a major problem, as well as moving in traffic. Someone also mentioned, that added to having related persons and friends there was a problem of misunderstanding even among those closest to them.

Many of the participants were active members in various types of associations regardless of their disabilities.

The sub-theme *Time perspective* with category Life history provides the basis for how one looks at one’s self-image and how to deal with one’s problems in life. The vision for future indicated some concern about further impaired hearing, the lack of independence, isolation, hopelessness, but also acceptance and gratitude for the little sight or hearing one has left.

Surprisingly, the Combination’s effects and Limitation in sub-theme *Disability*, demonstrated that most of the participants did not think of their vision and hearing impairment as a combined disability. One possible reason may be that the disabilities often progressed independent of each other. Demographic data shows that the hearing impairment existed in some cases from birth, some participants had the vision impairment when they were born, and most of the participants acquired disabilities in childhood or later in life. When a person is coping to one disability and the other is added and gets worse and worse, then it was perceived as a more problematic situation. Many participants declared the content *isolation* as a huge problem. The second content having *control* and influence *over one’s own life* promotes self-confidence and self-esteem. Contrary to this lack of control and influence over one’s own living conditions may result in feelings of exclusion and impotence [54]. The combined hearing and vision disabilities limit in all aspects of daily life e.g., reading, moving around, shopping, and participants announce that they need support in daily life activities. Persons with dual sensory loss are depending on a person by her/his side in many situations. Our results indicate that the deafblindness create even more isolation than having one disability, and that the greater the disabilities are the greater the challenges are.

Sub-theme *Challenges* harbours many categories. To avoid misunderstandings, there is a need of more information to the environment about disabilities. Participants highlights that dual sensory loss takes energy and that there is a sadness beneath the surface related to things you can no longer do. This study identified, similar to Hallberg [55] showed, the importance of accepting one’s disability and integrating it with one’s self-image in order to feel as valuable as other individuals.

In sub-theme *Needs* appeared the importance of continuity in health care and the positive routines at the Deafblind-team unit as they perform an annual appointment for every patient. It emerged that it is important with contacts with the surrounding world to avoid isolation and to gain accessibility in society. Radio and access to social media demonstrated an important area to be able to obtain information. All participants had various kinds of technical tools for their hearing and vision impairment. Another tool, the white cane, was discovered as the “most loyal companion”, but it took time for some participants to identify that. It turned out that the white cane implies two sides of a coin; help and stigma. Some participants said that they felt vulnerable in certain situations and did not want to show their surroundings that they had vision problems. At the same time, the white cane was very helpful. The Deafblind-team unit has white cane training, which the participants appreciate because the white cane is of great importance and should never be underestimated. The present study indicates similar findings as Ehn et al [53] that to accustom to the white cane takes time and that the experience of using it becomes more positive over time.

The companion service will promote equality in living conditions and full participation in society according to The Swedish National Board of Health and Wellfare (Socialstyrelsen) [40]. Unfortunately, the companion services support in Sweden has become much more difficult to receive with many limitations regarding the length of support time one can get. Offering a few hours of support per week for this patient group does not meet with the statement described above. Only three of the 14 participants came with companion service person to our interviews, and one was accompanied by a relative. To increase accessibility in the society, it is important to offer adequate opportunity for support through companion service, otherwise the requirements of the legislation are not met with this patient group.

The sub-theme *Coping strategies* showed a high degree of invention of the participants both at home and abroad. The study confirms that people strive to be considered as normal as possible, despite their disability, by using different coping strategies in similar ways as previous studies have shown [34, 55]. Many participants asked for support when they were outside home, and they used companion service when possible. One of them used a rope attached to a person in front of him/her and with help of the stretch of the rope, could walk or jog in different terrains or swim in the pool. One strategy could be to provide information to the public of the importance of presenting oneself every time one is near to a person with a vision impairment and not to speak from another room to a person with a hearing impairment.

The researchers’ preconceived notion was that the participants did not perceive their vision and hearing impairment as a combination. Many described more likely a separate vision and hearing impairment. Our “pre-understanding” was that patients might have various challenges due to their dual disability, which can cause various expressions. The interviews showed which kind of difficulties patients with dual sensory loss had, and with what kind of strategies they master these difficulties. In light of patient’s experiences, the findings showed even how positive, energetic, committed, and active participants were in the present study.

The origin of the health and the salutogenic theory by Antonovsky that emphasizes the human ability to cope with difficulties were present in all participants in the present study [56, 57].

The focus is not only on illness / functional ability but also on the ability to handle difficulties. This sense of coherence was observed in how participants managed their daily lives using many strategies.

### Strengths and limitations

The qualitative design was employed, and inductive content analysis was conducted as an appropriate technique to describe verbal phenomena [42]. The qualitative study with interviews describes and increases the understanding of the research questions and the reality being studied.

In qualitative research the trustworthiness includes credibility, transferability and dependability [45] and the study strengthened these by involving four authors in analysis of the meaning units and codes, categorisation and finally the main themes. Audio recorded interviews and authentic citations increase the trustworthiness of the study [41]. The researchers describes data collection and analysis process to facilitate transferability [45], and by representative quotations [45]. The present study used qualitative method with semi-structured open-ended questions. A qualitative research strategy opens up the possibilities for the researcher’s own bias. To reduce the researcher’s own bias in the study the transcribed text was read several times by the authors to reach consensus.

All interviews except one, were conducted in a hospital setting, which minimizes the source of error depending on varying environments.

In the study, both interviewers participated together in each meeting with the participants. Every other interview they changed acting roles and were either interviewer or observer. This was perceived as a strength to gain insight and knowledge about each interview situation. The observation notes, while one of the interviewers observed the other interviewer, reflected each interview, and the observations were discussed between them. This method was used in order to give feedback to each other, and this manner also improved the interview technique by time. The researchers did not feel that the participants were disturbed by this method because the other sat quiet during the interview after introducing herself to the participant.

Three of the participants did not return their consent form to quote verbatim in text. The researchers do not know if these three had understood the meaning of the request or if they really did not want their quote to be used which were sent by letter after the interviews were completed. This fact made the study unable to reproduce these three participants’ quotes.

Nevertheless, these quotes were interpreted and contributed to the study.

## Conclusions

One of the key words for the study is that the combination of vision and hearing impairment evokes isolation. The independence to spontaneous doing things and get outside door, is limited because they need support of other people in everyday life. At the same time, the participants show creativity, willingness to manage themselves, exhibit a positive life philosophy, see opportunities, and that most things can be performed but in a variety of ways. The experiences identified among participants, in the present study, are positive and they want to be as equal as everyone else, and to have control over their own lives.

The study indicates that it is the combination of dual loss that makes difficulties in daily life by causing, for example, isolation. How can health care professionals, meet the needs of this patient group? There is a great lack of understanding in society for this combination of vision and hearing impairment and therefore there is an urgent need of more information and education about dual sensory loss in the society.

One recommendation is that meeting an individual with a combined hearing and vision impairment should immediately alert the health care as these patients belong to a extremely vulnerable group. Furthermore, the community-organized companion services should be investigated and reviewed to meet with the needs of this patient group.

## Data Availability

All relevant data are within the manuscript and its Supporting Information files.

## Supporting information

S1 Appendix. Key informant interview guide. (PDF)

## Acknowledgments

We are grateful to all participants who took part in interviews in this study and shared their experiences, and to our colleagues who helped with recruitment of participants.

We also thank Adrian Taheri for transcription.

## References

1. Strawbridge WJ, Wallhagen MI, Shema SJ, Kaplan GA. Negative consequences of hearing impairment in old age: a longitudinal analysis. The Gerontologist. 2000;40(3):320–6. Epub 2000/06/15. doi: 10.1093/geront/40.3.320. PubMed PMID: 10853526.

2. GBD. Hearing loss prevalence and years lived with disability, 1990-2019: findings from the Global Burden of Disease Study 2019. Lancet (London, England). 2021;397(10278):996–1009. Epub 2021/03/15. doi: 10.1016/s0140-6736(21)00516-x. PubMed PMID: 33714390; PubMed Central PMCID: PMCPMC7960691.

3. Statistiska Centralbyrån. Undersökningar av levnadsförhållanden (ULF/SILC) 2015-2016 [2019 July 04]. Available from: http://www.statistikdatabasen.scb.se/pxweb/sv/ssd/STARTLELE0101LE0101H/LE0101H13/table/tableViewLayout1/.

4. Q-ORL. Swedish Quality Register of Otorhinolaryngology. Nationellt kvalitetsregister för öron-, näs-och halssjukvård. 1997 [2022 Mars 09]. Available from: https://www.entqualitysweden.se.

5. Löfvenberg C, Carlsson PI, Barrenäs ML, Skagerstrand Å, Simic D, Carlsson J, et al. Prevalence of severe-to-Profound hearing loss in the adult Swedish population and comparison with cochlear implantation rate. Acta oto-laryngologica. 2022:1–5. Epub 2022/06/01. doi: 10.1080/00016489.2022.2073388. PubMed PMID: 35635283.

6. Clark JG. Uses and abuses of hearing loss classification. Asha. 1981;23(7):493–500. Epub 1981/07/01. PubMed PMID: 7052898.

7. WHO. World Heath Organization. Grades of hearing impairment 1991 [2019 July 03]. Available from: https://apps.who.int/iris/bitstream/handle/10665/58839/WHO_PDH_91.1.pdf?sequence=1&isAllowed=y.

8. Desrosiers J, Wanet-Defalque MC, Temisjian K, Gresset J, Dubois MF, Renaud J, et al. Participation in daily activities and social roles of older adults with visual impairment. Disability and rehabilitation. 2009;31(15):1227–34. Epub 2009/10/06. PubMed PMID: 19802927.

9. Schilling OK, Wahl HW, Horowitz A, Reinhardt JP, Boerner K. The adaptation dynamics of chronic functional impairment: what we can learn from older adults with vision loss. Psychology and aging. 2011;26(1):203–13. Epub 2010/12/15. doi: 10.1037/a0021127. PubMed PMID: 21142375.

10. GBD. Trends in prevalence of blindness and distance and near vision impairment over 30 years: an analysis for the Global Burden of Disease Study. The Lancet Global health. 2021;9(2):e130–e43. Epub 2020/12/05. doi: 10.1016/s2214-109x(20)30425-3. PubMed PMID: 33275950; PubMed Central PMCID: PMCPMC7820390.

11. Swedish Eye Centres (Syncentralerna). Information about vision impairment 2019 [2021 May 06]. Available from: https://www.srf.nu/.

12. World Health Organization (WHO). The International Classification of Diseases-11. Blindness and vision impairment 2018 [2019 July 10]. Available from: https://www.who.int/en/news-room/fact-sheets/detail/blindness-and-visual-impairment.

13. World Health Organization (WHO). ICD-11 for Mortality and Morbidity Statistics. Version: 04/2019. Vision impairment including blindness [2019 July 10]. Available from: https://icd.who.int/browse11/l-m/en#/ http://id.who.int/icd/entity/1103667651.

14. Dalton DS, Cruickshanks KJ, Klein BE, Klein R, Wiley TL, Nondahl DM. The impact of hearing loss on quality of life in older adults. The Gerontologist. 2003;43(5):661–8. Epub 2003/10/23. PubMed PMID: 14570962.

15. Heine C, Browning CJ. Communication and psychosocial consequences of sensory loss in older adults: overview and rehabilitation directions. Disability and rehabilitation. 2002;24(15):763–73. Epub 2002/11/20. doi: 10.1080/09638280210129162. PubMed PMID: 12437862.

16. WFDB. World Federation of the Deafblind. At risk of exclusion from CRPD and SDGs implementation: inequality and persons with deafblindness. 2018.

17. Turunen-Taheri S, Skagerstrand A, Hellstrom S, Carlsson PI. Patients with severe-to-profound hearing impairment and simultaneous severe vision impairment: a quality-of-life study. Acta oto-laryngologica. 2017;137(3):279–85. Epub 2016/09/24. doi: 10.1080/00016489.2016.1229025. PubMed PMID: 27659206.

18. Förbundet Sveriges Dövblinda. Swedish Association of the Deafblind 2007, rev 2016 [2021 May 06]. Available from: http://www.fsdb.org/Nordisk-definitionavdovblindhet.html.

19. The Swedish National Information Centre for Deafblindness Issues. Nordic definition of deafblindness 2016 [2021 May 06]. Available from: https://nkcdb.se/dovblindhet/fakta-om-dovblindhet/nordisk-definition/.

20. Socialdepartementet. Statens offentliga utredningar. Teckenspråk och teckenspråkiga : översyn av teckenspråkets ställning : slutbetänkande Stockholm: Fritze; 2006 [2021 May 06]. Available from: https://www.regeringen.se/rattsliga-dokument/statens-offentliga-utredningar/2006/05/sou-200654/.

21. Lundin E, Widén SE, Wahlqvist M, Anderzén-Carlsson A, Granberg S. Prevalence, diagnoses and rehabilitation services related to severe dual sensory loss (DSL) in older persons: a cross-sectional study based on medical records. International journal of audiology. 2020;59(12):921–9. Epub 2020/07/07. doi: 10.1080/14992027.2020.1783003. PubMed PMID: 32628050.

22. National Knowledge Centre for Deafblind issues. Nationellt kunskapscenter för Dövblindfrågor. Prevalence of deafblindness 2018 [2021 May 06]. Available from: https://nkcdb.se/dovblindhet/fakta-om-dovblindhet/forekomst/.

23. Sadeghi AM, Cohn ES, Kimberling WJ, Halvarsson G, Moller C. Expressivity of hearing loss in cases with Usher syndrome type IIA. International journal of audiology. 2013;52(12):832–7. Epub 2013/10/29. doi: 10.3109/14992027.2013.839885. PubMed PMID: 24160897.

24. Mathur P, Yang J. Usher syndrome: Hearing loss, retinal degeneration and associated abnormalities. Biochimica et biophysica acta. 2015;1852(3):406–20. Epub 2014/12/08. doi: 10.1016/j.bbadis.2014.11.020. PubMed PMID: 25481835; PubMed Central PMCID: PMCPmc4312720.

25. Sadeghi M, Kimberling W, Tranebjœrg L, Möller C. The prevalence of Usher Syndrome in Sweden: a nationwide epidemiological and clinical survey. Audiological Medicine. 2004;2(4):220–8. doi: 10.1080/16513860410003093.

26. Brannstrom KJ, Basjo S, Larsson J, Lood S, Lunda S, Notsten M, et al. Psychosocial work environment among Swedish audiologists. International journal of audiology. 2013;52(3):151–61. Epub 2012/12/12. doi: 10.3109/14992027.2012.743045. PubMed PMID: 23216266.

27. Optician Association. Optikerförbundet 2021 [2021 May 06]. Available from: https://optikerforbundet.se/.

28. Förbundet Sveriges Dövblinda. Swedish Association of the Deafblindness. Dövblindteam 2019 [2019 July 11]. Available from: http://www.fsdb.org/doevblindteam.html.

29. MFD. Swedish Agengy for Participation. Myndigheten för delaktighet (MFD). The UN Convention on the Rights of Persons with Disabilities. FN:s konvention om rättigheter för personer med funktionsnedsättning. 2009 [2021 May 06]. Available from: https://www.mfd.se/kunskap/ and http://www.mfd.se/delaktighet/fns-konvention/.

30. Regeringskansliet. Government Offices of Sweden. The UN Convention on the Rights of Persons with Disabilities. SÖ 2008:26. Regeringskansliet. Konvention om rättigheter för personer med funktionsnedsättning och fakultativt protokoll. 2008 [2021 May 06]. Available from: https://www.regeringen.se/informationsmaterial/2015/06/konvention-om-rattigheter-for-personer-med-funktionsnedsattning/.

31. Joge Johansson C, Edberg P-O., & Nylander G. Plus & Minus. Framgångsfaktorer och hindrande faktorer i samband med förvärvad dövblindhet. 2010 ISBN: 978-91-978034-5-8. https://nkcdb.se/wp-content/uploads/2015/02/Plus-och-Minus.pdf.

32. Swedish Agency for Accessible Media (MTM). Myndigheten för tillgängliga medier. 2019 [2021 May 06]. Available from: https://www.mtm.se/.

33. Ringdahl A, Grimby A. Severe-profound hearing impairment and health-related quality of life among post-lingual deafened Swedish adults. Scandinavian audiology. 2000;29(4):266–75. Epub 2001/02/24. PubMed PMID: 11195947.

34. Backenroth GAM, Ahlner BH. Quality of life of hearing-impaired persons who have participated in audiological rehabilitation counselling. International Journal for the Advancement of Counselling. 2000;22(3):225–40. doi: 10.1023/A:1005655017175.

35. Turunen-Taheri S, Carlsson PI, Johnson AC, Hellstrom S. Severe-to-profound hearing impairment: demographic data, gender differences and benefits of audiological rehabilitation. Disability and rehabilitation. 2019;41(23):2766–74. Epub 2018/06/13. doi: 10.1080/09638288.2018.1477208. PubMed PMID: 29893149.

36. Ehn M, Anderzén-Carlsson A, Möller C, Wahlqvist M. Life strategies of people with deafblindness due to Usher syndrome type 2a - a qualitative study. International journal of qualitative studies on health and well-being. 2019;14(1):1656790. Epub 2019/09/01. doi: 10.1080/17482631.2019.1656790. PubMed PMID: 31470768; PubMed Central PMCID: PMCPMC6735326.

37. Synskadades Riksförbund. Vardagstips för personer med synnedsättning. The National Association of the Visually Impaired 2017 [2021 May 06]. Available from: https://www.srf.nu/globalassets/informationsmaterial/for-medlemmar-och-andra-synskadade/vardagstips_2017_tillganglig.pdf.

38. Sveriges Riksdag. The Social Services Act. Socialtjänstlag (2001:453) 2001 [2021 May 06]. Available from: https://www.riksdagen.se/sv/dokument-lagar/dokument/svensk-forfattningssamling/socialtjanstlag-2001453_sfs-2001-453.

39. Stockholms Stad. Companion service for persons with disability. Ledsagning för personer med funktionsnedsättning. 2021 [2021 May 06]. Available from: https://www.stockholm.se/FamiljOmsorg/funktionsnedsattning/Personligt-stod/Ledsagning/.

40. Socialstyrelsen. Socialstyrelsen. The Swedish National Board of Health and Welfare. Ledsagning enligt LSS och SoL. 2011 [2021 May 06]. Available from: https://www.socialstyrelsen.se/globalassets/sharepoint-dokument/artikelkatalog/ovrigt/2011-3-49.pdf.

41. Patton MQ. Qualitative Research and Evaluation Methods. Thousand Oaks, CA: SAGE Publications; 2014.

42. Krippendorff K. Content analysis : an introduction to its methodology. Thousand Oaks, Calif. ;: SAGE; 2013.

43. Elo S, Kyngas H. The qualitative content analysis process. Journal of advanced nursing. 2008;62(1):107–15. Epub 2008/03/21. doi: 10.1111/j.1365-2648.2007.04569.x. PubMed PMID: 18352969.

44. Erlingsson C, Brysiewicz P. A hands-on guide to doing content analysis. African journal of emergency medicine : Revue africaine de la medecine d’urgence. 2017;7(3):93–9. Epub 2017/09/01. doi: 10.1016/j.afjem.2017.08.001. PubMed PMID: 30456117; PubMed Central PMCID: PMCPmc6234169.

45. Graneheim UH, Lundman B. Qualitative content analysis in nursing research: concepts, procedures and measures to achieve trustworthiness. Nurse education today. 2004;24(2):105–12. Epub 2004/02/11. doi: 10.1016/j.nedt.2003.10.001. PubMed PMID: 14769454.

46. Polit DF, Beck CT. Generalization in quantitative and qualitative research: myths and strategies. International journal of nursing studies. 2010;47(11):1451–8. Epub 2010/07/06. doi: 10.1016/j.ijnurstu.2010.06.004. PubMed PMID: 20598692.

47. Malterud K. Qualitative research: standards, challenges, and guidelines. Lancet (London, England). 2001;358(9280):483–8. Epub 2001/08/22. doi: 10.1016/s0140-6736(01)05627-6. PubMed PMID: 11513933.

48. Cope DG. Methods and meanings: credibility and trustworthiness of qualitative research. Oncology nursing forum. 2014;41(1):89–91. Epub 2013/12/26. doi: 10.1188/14.onf.89-91. PubMed PMID: 24368242.

49. Braun V, Clarke V. Using thematic analysis in psychology. Qualitative Research in Psychology. 2006;3(2):77–101. doi: 10.1191/1478088706qp063oa.

50. O’Brien BC, Harris IB, Beckman TJ, Reed DA, Cook DA. Standards for reporting qualitative research: a synthesis of recommendations. Academic medicine : journal of the Association of American Medical Colleges. 2014;89(9):1245–51. Epub 2014/07/01. doi: 10.1097/acm.0000000000000388. PubMed PMID: 24979285.

51. Tong A, Sainsbury P, Craig J. Consolidated criteria for reporting qualitative research (COREQ): a 32-item checklist for interviews and focus groups. International journal for quality in health care : journal of the International Society for Quality in Health Care. 2007;19(6):349–57. Epub 2007/09/18. doi: 10.1093/intqhc/mzm042. PubMed PMID: 17872937.

52. Wahlqvist M, Moller C, Moller K, Danermark B. Physical and Psychological Health in Persons with Deafblindness That Is Due to Usher Syndrome Type II. Journal of Visual Impairment & Blindness. 2013;107(3):207-20. PubMed PMID: WOS:000209257700005.

53. Ehn M, Anderzen-Carlsson A, Moller C, Wahlqvist M. Life strategies of people with deafblindness due to Usher syndrome type 2a - a qualitative study. 2019;14(1):1656790. doi: 10.1080/17482631.2019.1656790. PubMed PMID: 31470768.

54. Regeringskansliet. Government Offices of Sweden. Good and equal health - a developed one health policy. Proposition 2017/18:249. Regeringskansliet. God och jämlik hälsa – en utvecklad folkhälsopolitik 2017 [2019 July 12]. Available from: https://www.regeringen.se/498282/contentassets/8d6fca158ec0498491f21f7c1cb2fe6d/prop.-2017_18_249-god-och-jamlik-halsa--en-utvecklad-folkhalsopolitik.pdf.

55. Hallberg LR-M. Stigmatisering, coping och handikappupplevelse: Intervjuer med medelålders personer med fórvärvade hörselskador. Nordic Journal of Nursing Research. 1996;16:39 – 45.

56. Antonovsky A. Health, stress, and coping. San Francisco: Jossey-Bass; 1979.

57. Antonovsky A. Unraveling the mystery of health: How people manage stress and stay well. San Francisco, CA, US: Jossey-Bass; 1987. xx, 218-xx, p.

